# Maternal Vaginal Pathogenic Bacterial Colonization and Early-Onset Neonatal Sepsis in Mwanza City, Tanzania: A Prospective Cohort Study

**DOI:** 10.64898/2026.10.22.26366266

**Authors:** Dismas Matovelo, Paul Alikado Sabuni, Godfrey W Kajungu, Michael James, Benson Kidenya, Moke Magoma, Jennifer A. Downs, Jeremiah Seni

**Affiliations:** Catholic University of Health and Allied Sciences, Weill Bugando School of Medicine, Department of Obstetrics & Gynecology, Mwanza, Tanzania; Catholic University of Health and Allied Sciences, School of Public Health, Department of Epidemiology & Biostatistics, Mwanza, Tanzania; Catholic University of Health and Allied Sciences, Weill Bugando School of Medicine, Department of Microbiology & Immunology, Mwanza, Tanzania; Engender Health, Tanzania; Catholic University of Health and Allied Sciences, Weill Bugando School of Medicine, Department of Medicine, Mwanza, Tanzania; Center for Global Health, Weill Cornell Medicine, New York, USA

**Keywords:** Early-onset neonatal sepsis, vaginal pathogenic bacterial colonization, HIV, Tanzania

## Abstract

**Background:** Early-onset neonatal sepsis (EONS) remains a major cause of neonatal morbidity and mortality in low- and middle-income countries. Maternal vaginal pathogenic bacterial colonization and HIV infection may increase the risk of EONS through vertical transmission and immune dysregulation, but prospective evidence from sub-Saharan Africa is limited. This study assessed the association between maternal HIV status, vaginal pathogenic bacterial colonization, and EONS among women delivering at five selected health facilities in Mwanza City, Tanzania.

**Methods:** A prospective comparative cohort study was conducted between August 2023 and February 2025 among 580 pregnant women delivering at five selected health facilities in Mwanza City. A maternal lower vaginal swab was collected before delivery for bacterial culture and antimicrobial susceptibility testing. Neonates were followed for seven days after birth to identify EONS using standardized clinical and laboratory criteria, with culture confirmation where applicable. Kaplan–Meier survival analysis and Cox proportional hazards regression were used to estimate incidence rates and identify independent predictors of neonatal sepsis.

**Results:** Overall, 89 neonates developed EONS, yielding a cumulative incidence of 15.3% and an incidence rate of 2.42 per 100 person-days (95% CI: 1.97–2.98). Twenty-four (27.0%) cases were culture-confirmed, while 65 (73.0%) were clinically diagnosed. Neonates born to colonized mothers had substantially higher sepsis incidence than those born to non-colonized mothers (3.41 vs. 0.34 per 100 person-days). After adjustment, maternal vaginal bacterial colonization was the strongest predictor of neonatal sepsis (adjusted hazard ratio [aHR] 9.52, 95% CI: 3.47–26.09; p<0.001). Maternal HIV infection independently increased the hazard of neonatal sepsis by 76% (aHR 1.76, 95% CI: 1.14–2.73; p=0.011), while maternal malaria doubled the risk (aHR 2.74, 95% CI: 1.43–5.22; p=0.002). *Escherichia coli* (*E. coli*) accounted for 58.3% of culture-confirmed neonatal infections, followed by both *Klebsiella pneumoniae* and *Staphylococcus aureus* (20.8% each).

**Conclusion:** Maternal vaginal pathogenic bacterial colonization was strongly associated with early-onset neonatal sepsis. Maternal HIV infection and malaria were also independently associated with increased sepsis. The findings support strengthening maternal infection prevention, neonatal surveillance, and microbiological diagnostic capacity, while prospective intervention studies are needed to determine whether targeted screening and prevention of maternal bacterial colonization can reduce neonatal sepsis.

## INTRODUCTION

Early-onset neonatal sepsis (EONS) remains a leading cause of neonatal morbidity and mortality globally, largely affecting low- and middle-income countries (LMICs), particularly in the sub-Saharan Africa (SSA). Despite advances in neonatal care, sepsis continues to account for a substantial proportion of neonatal deaths, with case fatality rates in LMICs ranging from 10-30% (1). In addition to mortality, survivors are at increased risk of long-term neurodevelopmental impairment, underscoring the critical need for improved prevention strategies. Recent systematic reviews report an overall pooled magnitude of neonatal sepsis in SSA of 36% to 41% (2–4), while early sepsis in Tanzania ranges from 11% to 16% (5, 6).

The pathogenesis of EONS is closely linked to vertical transmission of pathogenic microorganisms from the maternal genital tract during labour and delivery, as well as exposure to contaminated environments. Maternal vaginal bacterial colonization plays a pivotal role in this process and has been associated with a spectrum of adverse maternal and neonatal outcomes, including neonatal sepsis, postpartum endometritis, urinary tract infections, and chorioamnionitis(7). Common pathogens such as *Escherichia coli*, *Klebsiella pneumoniae*, *Group B Streptococcus, Staphylococcus aureus*, and *Acinetobacter* species are frequently implicated, underscoring the importance of maternal microbiological profiles in shaping neonatal infection risk (7, 8). A previous study of neonatal sepsis in Ethiopia found that maternal bacterial colonization contributed to 37.4% of cases (8).

Maternal HIV infection may further modify the risk of neonatal sepsis through complex immunological and microbiological mechanisms. Infants born to pregnant women living with HIV (PWLHIV), including those who are HIV-exposed but uninfected, have been shown to experience higher rates of infectious morbidity. This increased susceptibility has been attributed to altered maternal immunity, in utero immune modulation, and potential disruptions in maternal–fetal microbial transfer (9, 10). However, existing evidence remains inconsistent, and the relationships between maternal HIV status, microbiome alterations, and neonatal sepsis remain unclear (11–13). These discrepancies suggest that contextual factors, including maternal bacterial colonization patterns, antimicrobial exposure, and health system differences, may play a critical role in modifying this relationship (12).

In SSA, and particularly in Tanzania, there is a paucity of prospective data examining the interplay between maternal vaginal bacterial colonization, HIV infection, and neonatal sepsis. Most available studies are cross-sectional, limiting causal inference, and few include longitudinal follow-up of neonates to establish temporal relationships between maternal exposures and neonatal outcomes. Furthermore, microbiological characterization of colonizing and infecting organisms, including antimicrobial resistance (AMR) profiles, remains underexplored despite growing concerns about rising AMR in neonatal sepsis management (14, 15).

This study investigated the association between maternal HIV status, vaginal pathogenic bacterial colonization, and early-onset neonatal sepsis among women delivering at five selected health facilities in Mwanza City, Tanzania. By integrating prospective clinical and microbiological data and following neonates for the first 7 days after birth, the study sought to generate context-specific evidence to inform targeted interventions to prevent and manage neonatal sepsis in Tanzania and other similar LMICs.

## METHODS

### Study design and settings

A comparative cohort study conducted from August 2023 to February 2025 in five selected high-volume labour wards in Mwanza City, namely Bugando Medical Centre (BMC), Sekou Toure Regional Referral Hospital (STRRH), Nyamagana District Hospital (NDH), Buzuruga Health Centre (BHC), and Igoma Health Centre (IHC) which account for approximately 53% (2,100/4,000) of all institutional deliveries in the City. However, BMC serves as a major zonal referral hospital for 16 million people with a 950-bed capacity; STRRH covers a population of 2.7 million with a 350-bed capacity; and NDH, BHC, and IHC support both Nyamagana and Ilemela districts within Mwanza City (catchment population of 1.2 million).

### Study population

Pregnant women delivering at five selected health facilities, and neonates born to these women, were followed up to seven days from birth to assess the occurrence of neonatal sepsis. Inclusion criteria comprised a gestational age of ≥28 weeks in the latent phase of labour (those who had uterine contractions, cervical effacement, and dilatation of up to 5cm) and were planned for delivery. Exclusion criteria included pregnant women with multiple pregnancies, vaginal bleeding, those with cervical cerclage in situ, and those referred from elsewhere to deliver in these facilities.

### Sample size and sampling procedure

The study sample size was calculated using a double-proportional formula (16), primarily powered to compare maternal vaginal bacterial colonization between (PWLHIV) and HIV-uninfected pregnant women (HUPW), based on previously reported Group B Streptococcus colonization rates of 24.0% among PWLHIV and 13.7% among HUPW(17), with a two-sided α of 0.05, 80% power, and a 1:2 allocation ratio, the minimum required sample size was 522 participants (174 (PWLHIV) and 348 HUPW). The additional 10% to added to account for loss to follow-up and maximize the sample size; therefore, we continued recruitment until 580 women were enrolled.

Based on published studies in Tanzania, we anticipated an incidence of early neonatal sepsis of approximately 16% among neonates (5, 6), in which the recruited cohort would yield sufficient neonatal sepsis events to compare sepsis rates between neonates born to women with and without HIV and to perform multivariable Cox proportional hazards regression analyses. This neonatal cohort analysis was pre-specified within a larger prospective study investigating maternal vaginal bacterial colonization. We used consecutive sampling to recruit eligible patients until we reached the target sample size. The sample size for each facility was based on the proportional allocation formula.

### Data collection

Upon admission to the maternity ward, ANC cards were reviewed to determine patient eligibility and to confirm gestational age via first-trimester ultrasound, reliable LNMP, or second-trimester ultrasound. Women with documented HIV-negative status from prior ANC visits were counselled for re-testing, and a comprehensive general and obstetrical examination was conducted on all participants.

Socio-demographic and clinical characteristics, as well as personal behaviours, were also collected using a pre-tested, structured questionnaire developed for the study, informed by ANC guidelines and previous studies on vaginal bacterial colonization (S1 Appendix). Additional clinical conditions likely to confound the association were assessed at enrolment, including a blood smear for malaria, random blood glucose for diabetes mellitus, urinalysis for UTI and a rapid point-of-care test [POC-CCA3, Mondial Diagnostics, Netherlands] for schistosomiasis. Haemoglobin levels were also tested to assess for anemia, and blood pressure measurements for hypertensive disorders of pregnancy (HDP).

Prior to delivery, trained personnel collected a lower vaginal swab from asymptomatic pregnant women without using a speculum. A sterile, pre-packaged swab containing Amies Agar Gel without charcoal (Copan Diagnostics Inc., Murrieta, CA) was inserted into the vagina halfway between the introitus and the cervix. The swab was gently pressed against the vaginal walls (2–3 cm deep) and rotated for at least 30 seconds. To prevent contamination, the labia were held apart while the swab was carefully extracted without touching the skin or anal region. The vaginal swabs with pre-packed Amies agar were immediately placed in the Amies transport medium in a specimen transport ice cooler at 4°C, and blood samples for neonates with features of neonatal sepsis were taken to the CUHAS Multipurpose Laboratory within 2 hours for microbiological analysis. Culture was performed for each sample using standard guidelines on Blood agar, MacConkey agar (OXOID, UK), and CHROMagarStreB^TM^, and incubated at 35 to 37°C for 18 to 24 hours.

Because the vagina is a non-sterile site that naturally hosts diverse microbiota, we applied strict criteria to determine whether bacterial isolates were eligible for biochemical identification and antimicrobial susceptibility testing (AST). We further processed isolates that met two conditions: first, significant culture growth yielding ≥ 10^5^ colony-forming units (CFUs) per millilitre for a single bacterial morphotype, and second, growth extending into at least two quadrants of the culture plate. We used standard biochemical methods to identify both Gram-positive and Gram-negative bacteria (18). The antimicrobial susceptibility testing was performed on all identified bacteria using the conventional Kirby-Bauer disk diffusion method on Mueller-Hinton agar (Oxoid, UK). The zones of inhibition or diameters were interpreted as resistant, intermediate, or sensitive using the Clinical Laboratory Standards Institute (CLSI) guidelines for specific management, and the detection of antimicrobial resistance phenotypes was based on the double-Disk Synergy Test (19).

### Variable Measurement

For this study, vaginal pathogenic bacterial colonization was defined as the recovery of viable potentially pathogenic bacterial species from the lower vaginal mucosa of asymptomatic pregnant women using culture, with the organisms of interest selected on the basis of their recognized clinical relevance and alignment with bacterial groups represented in the WHO Bacterial Priority Pathogens framework (20). The EONS was defined as neonatal sepsis occurring from birth through day 7 of life using a modified framework that combines World Health Organization (WHO) clinical danger signs with laboratory criteria (21, 22). Clinical inclusion required at least one clinical danger sign: temperature instability (fever >38°C or hypothermia <35.5°C), feeding difficulties, lethargy, unconsciousness, convulsions, severe chest indrawing, or a respiratory rate ≥60 breaths/min. Laboratory criteria required elevated C-reactive protein (CRP >10 mg/L) and an abnormal leukocyte count (WBC <5,000/mm³ or >30,000/mm³) for symptomatic neonates ± culture isolation. Cases were stratified as either culture-confirmed sepsis if a recognized pathogen was isolated by blood culture, or as clinically suspected sepsis if cultures were negative. In this study, maternal sepsis was defined as a bacterial infection of the genital tract up to 7 days postpartum in which two or more of the following are present: pelvic pain, fever (temperature ≥ 38.5 °C on any occasion), foul-smelling lochia, and delayed uterine involution.

Predictor variables comprised demographic and clinical characteristics of mothers and neonates, alongside maternal factors such as HIV status, colonization status, and the presence of sepsis, to determine their effect on the incidence of neonatal sepsis.

### Data Management and Analysis

Data were initially managed and cleaned using Microsoft Excel before being imported into STATA version 18 for statistical analysis. Both descriptive and inferential analyses were performed. In the descriptive analysis, we used proportions to summarize categorical variables and means (with standard deviations) and medians (with interquartile ranges) to summarize continuous variables. Kaplan–Meier survival curves were applied to describe neonatal sepsis across categories of maternal HIV status, colonization status, and maternal sepsis, with the log-rank test used to compare incidence between groups. For inferential analysis, stepwise backward Cox proportional hazards regression models were used to control for confounders based on biological plausibility, and reported hazard ratios (HRs) with 95% confidence intervals (CIs) to indicate precision. We tested the proportional hazards (PH) assumption to assess the model’s adequacy. We conducted all statistical tests at the 5% significance level. Neonates who did not develop sepsis were censored at the date of their last documented follow-up.

### Ethical considerations

Ethical approvals were obtained from CUHAS/BMC (CREC/640/2023), NIMR (MR/53/100/731), and Weill Cornell Medicine (26-01029754-02), along with administrative permissions from regional and district health authorities. All participants provided written informed consent (thumbprint for illiterate participants) with guaranteed confidentiality and voluntary participation. Neonates with confirmed/clinical sepsis received IV broad-spectrum antibiotics, and participants with other health conditions or incidental findings were referred for standard care.

## RESULTS

### Demographic characteristics of the pregnant women

The median maternal age was 29.5 years (IQR 25–34), and most participants were aged 25–35 years (62.2%, n=361). Most were married (83.8%, n=486), and primary education was the most common education level (39.0%, n=226). Most women were entrepreneurs (56.0%, n=325) and paid out of pocket for health services (82.9%, n=481). A total of 89 neonates experienced sepsis within 7 days of birth. Among these, 27% (n=24) had culture-confirmed sepsis, and 73% (n=65) had clinically diagnosed sepsis (raised CRP and leucocytosis). About 2.2% (13) were not followed up over the full 7 days because they missed their postnatal care visit and could not be reached by phone. Residence was significantly associated with neonatal sepsis, with higher rates among those living outside Mwanza City than within the city (26.4% vs 13.8%, p=0.005) (S1 Table).

**S1 Table:** Demographic characteristics of the pregnant women stratified by neonatal sepsis (N=580).

| Variables | N (%) | Neonatal sepsis |  |  |
| --- | --- | --- | --- | --- |
| | | No (N=491)<br>n (%) | Yes (N=89)<br>n (%) | $\chi^2$ /t-test<br>(p-value) |
| **Maternal age (median, IQR)** | 29.5, 25-34 | 29, 25-34 | 30, 27-35 | 0.13 |
| **Age (in years)** |  |  |  | 0.84 |
| 17-24 | 123(21.2) | 106(86.2) | 17(13.8) |  |
| 25-35 | 361(62.2) | 305(84.5) | 56(15.5) |  |
| 36-44 | 96(16.6) | 80(83.3) | 16(16.7) |  |
| **Marital status** |  |  |  | 0.42 |
| Not married | 94(16.2) | 77(81.2) | 17(18.1) |  |
| Married | 486(83.8) | 414(85.2) | 72(14.8) |  |
| **Education level** |  |  |  | 0.18 |
| Primary education | 226(39.0) | 184(81.4) | 42(18.6) |  |
| Secondary education | 218(37.6) | 187(85.8) | 31(14.2) |  |
| College and above | 136(23.4) | 120(88.2) | 16(11.8) |  |
| **Occupation** |  |  |  | 0.66 |
| Not employed | 160(27.6) | 133(83.1) | 27(16.9) |  |
| Employed | 95(16.4) | 83(87.4) | 12(12.6) |  |
| Entrepreneur | 325(56.0) | 275(84.6) | 50(15.4) |  |
| **Practice douching** |  |  |  | 0.61 |
| No | 314(54.1) | 268(85.4) | 46(14.7) |  |
| Yes | 266(45.9) | 223(83.8) | 43(16.2) |  |
| **Body mass index (mean, SD)** | 25.02, 1.3 | 25.2(11.0) | 24.2(5.2) | 0.39 |
| **BMI** |  |  |  | 0.38 |
| Normal | 309(53.3) | 264(85.4) | 45(14.6) |  |
| Underweight | 35(6.0) | 26(74.3) | 9(25.7) |  |
| Overweight | 153(26.4) | 130(85.0) | 23(15.0) |  |
| Obesity | 83(14.3) | 71(85.5) | 12(14.5) |  |
| **Weight gain (mean, SD)** | 7.3, 4.9 | 7.3(4.9) | 7.0(4.0) | 0.51 |
| **Weight gain** |  |  |  | 0.28 |
| Too little (<8kg) | 359(61.9) | 306(85.2) | 53(14.8) |  |
| Normal (8kg – 12 kg) | 155(26.7) | 126(81.3) | 29(18.7) |  |
| Too much (>12kg) | 66(11.4) | 59(89.4) | 7(10.6) |  |
| **Frequency of sex per week (mean, SD)** | 2.4, 1.9 | 2.4(1.9) | 2.3(1.2) | 0.56 |
| **Sex frequency per week** |  |  |  | 0.33 |
| None | 28(4.8) | 27(96.4) | 1(3.6) |  |
| Once | 135(23.3) | 110(81.5) | 25(18.5) |  |
| 2 times | 170(29.3) | 145(85.3) | 25(14.7) |  |
| 3 times | 143(24.7) | 119(83.2) | 24(16.8) |  |
| 4 times and above | 104(17.9) | 90(86.5) | 14(13.5) |  |
| **Healthcare facility** |  |  |  | 0.27 |
| Bugando Medical Centre | 337(58.1) | 289(85.8) | 48(14.2) |  |
| Buzuruga Healthcare Centre | 46(7.9) | 40(87.0) | 6(13.0) |  |
| Igoma Healthcare Centre | 48(8.3) | 41(85.4) | 7(14.6) |  |
| Nyamagana District Hospital | 54(9.3) | 40(74.1) | 14(25.9) |  |
| Sekou Toure Regional Hospital | 95(16.4) | 81(85.3) | 14(14.7) |  |
| **Area of residence** |  |  |  | 0.005 |
| Outside Mwanza City | 72(12.4) | 53(73.6) | 19(26.4) |  |
| Within Mwanza City | 508(87.6) | 438(86.2) | 70(13.8) |  |

### Clinical characteristics of the pregnant women

Neonatal sepsis occurred more frequently among neonates born to mothers with malaria at enrolment (35.5% vs 14.2%, p=0.001) and PWLHIV (23.2% vs 11.3%, p<0.001). Colonization status was strongly associated with neonatal sepsis, occurring in 20.8% of neonates born to colonized mothers compared with 2.3% of those born to non-colonized mothers (p<0.001). Of 24 mother-neonate pairs with culture-confirmed neonatal sepsis, 91.7% (n=22) of isolates showed phenotypic concordance, while only 8.3% (n=2) did not. Neonates born to mothers with maternal sepsis also had a higher proportion of neonatal sepsis (34.9% vs 13.8%, p<0.001) (S2 Table).

**S2 Table:**
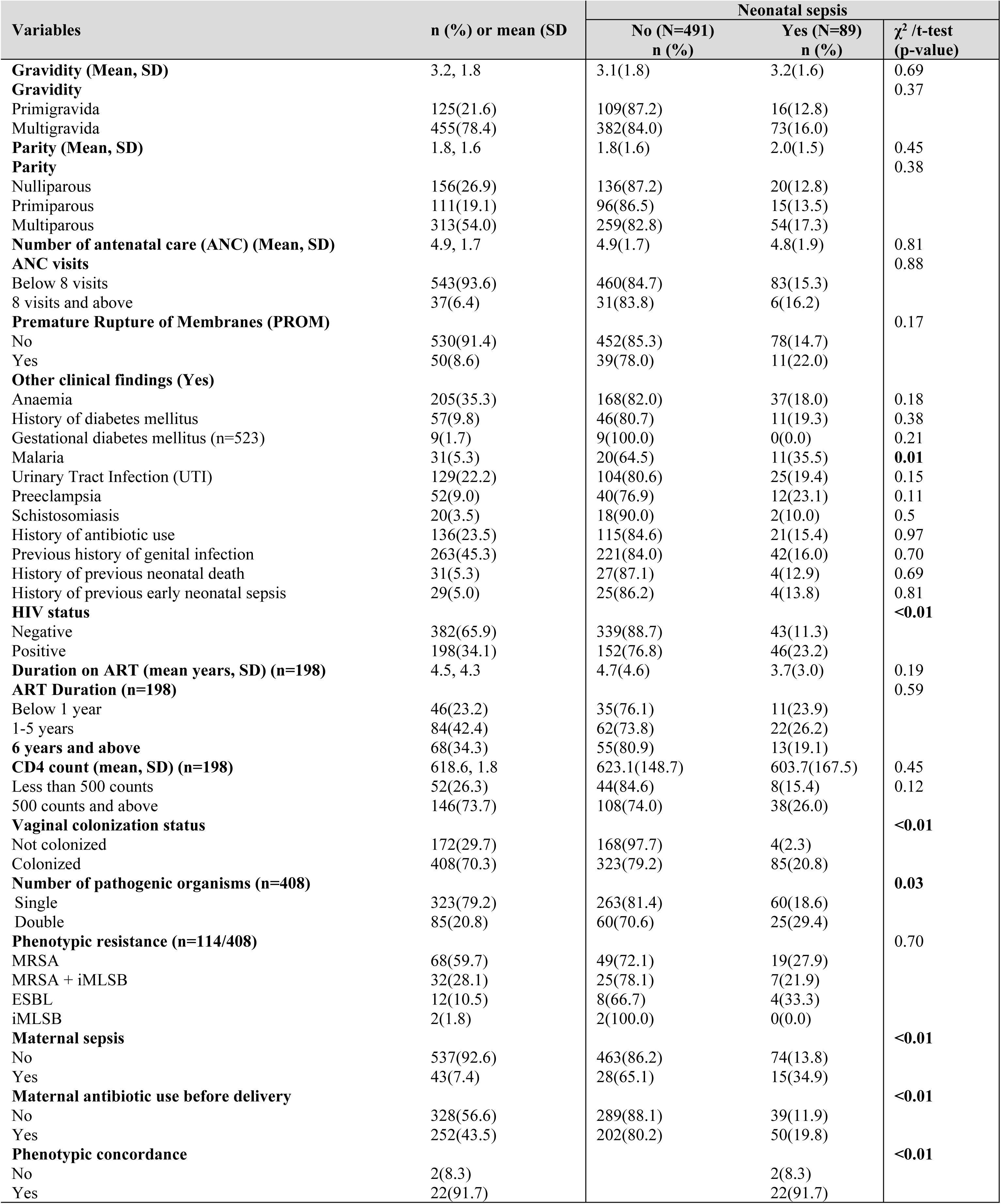
Clinical characteristics of the pregnant women stratified by neonatal sepsis (N=580).

Low birthweight (LBW <2.5 kg) occurred in 12.9% (n=75) of neonates and significantly increased the risk of neonatal sepsis to 25.3% (n=19; p=0.01), with *E. coli* (58.3%, n=14) as the most common bacterial isolate. Very LBW (<1.4 kg) neonates had the highest sepsis rate (62.5%; n=5; p=0.01) (S3 Table).

**S3 Table:** Intrapartum & Neonatal characteristics stratified by neonatal sepsis (N=580).

| Variables | n (%) or mean (SD) | Neonatal sepsis status |  |  |
| --- | --- | --- | --- | --- |
| | | No<br>n (%) | Yes<br>n (%) | $\chi^2$ /t-test<br>(p-value) |
| **Duration of labour in hours (Mean, SD)** | 8.6(4.3) | 8.6(4.3) | 8.0(4.7) | 0.18 |
| **Duration of rupture of membrane in hours (Mean, SD)** | 6.0(10.2) | 5.9(9.4) | 6.9(13.9) | 0.37 |
| **Number of vaginal Examinations (Mean, SD)** | 2.8(1.5) | 2.8(1.5) | 2.7(1.5) | 0.27 |
| **Mode of delivery** |  |  |  | 0.11 |
| Spontaneous vaginal delivery | 400(69.0) | 345(86.2) | 55(13.8) |  |
| Caesarean section | 180(31.0) | 146(81.1) | 34(18.9) |  |
| **Type of Caesarean section (n=180)** |  |  |  | 0.73 |
| Elective | 59(32.8) | 47(79.7) | 12(20.3) |  |
| Emergency | 121(67.2) | 99(81.8) | 22(18.2) |  |
| **Sepsis status (n=89)** |  |  |  |  |
| Clinical-diagnosed | 65(73.0) |  | 65(73.0) |  |
| Culture-confirmed | 24(27.0) |  | 24(27.0) |  |
| **Bacterial isolates (n=24)** |  |  |  |  |
| *E. coli* | 14(58.3) |  | 14(58.3) |  |
| *K. pneumoniae* | 5(20.8) |  | 5(20.8) |  |
| *S. aureus* | 5(20.8) |  | 5(20.7) |  |
| **Birth-weight (Mean, SD)** | 3.0(0.5) | 3.0(0.5) | 2.9(0.7) | **0.01** |
| **Birth-weight** |  |  |  | **0.01** |
| Normal birthweight ($\geq 2.5$kg) | 505(87.1) | 435(86.1) | 70(13.9) | |
| LBW (<2.5kg) | 75(12.9) | 56(74.7) | 19(25.3) |  |
| **LBW (n=75)** |  |  |  | **0.01** |
| Low (1.5kg-2.4kg) | 67(89.4) | 53(79.1) | 14(20.9) |  |
| Very Low (<1.4kg) | 8(10.6) | 3(37.5) | 5(62.5) |  |
| **Gestational age at birth (Mean, SD)** | 38.4(2.3) | 38.4(2.3) | 37.9(2.6) | 0.08 |
| **Gestational age at birth** |  |  |  | 0.27 |
| Post-term (Above 42 weeks) | 19(3.3) | 17(89.5) | 2(10.5) |  |
| Term (37-42 weeks) | 475(81.9) | 406(85.5) | 69(14.5) |  |
| Premature (<37 weeks) | 86(14.8) | 69(79.1) | 18(20.9) |  |
| **Premature birth (n=86)** |  |  |  | 0.59 |
| Premature late term (34-36 weeks) | 64(74.4) | 52(81.3) | 12(18.8) |  |
| Premature moderate term (32-33 weeks) | 9(10.5) | 6(66.7) | 3(33.3) |  |
| Premature very preterm (28-32 weeks) | 13(15.1) | 10(76.9) | 3(23.1) |  |

### The incidence of early neonatal sepsis among newborns

During 3,678 person-days of follow-up, the cumulative incidence was 15.3% (n=89), corresponding to an incidence rate of 2.42 per 100 person-days (95% CI: 1.97–2.98). Also, neonates born to non-colonized mothers had a very low incidence of 0.34 per 100 person-days (95% CI: 0.16–0.90), whereas those born to colonized mothers had a markedly higher incidence of 3.41 per 100 person-days (95% CI: 2.76–4.22). Furthermore, neonates born to HUPW had an incidence of 1.73 per 100 person-days (95% CI: 1.28–2.33), whereas those born to PWLHIV had more than double the incidence, at 3.86 per 100 person-days (95% CI: 2.89–5.15) (S4 Table).

**S4 Table:**
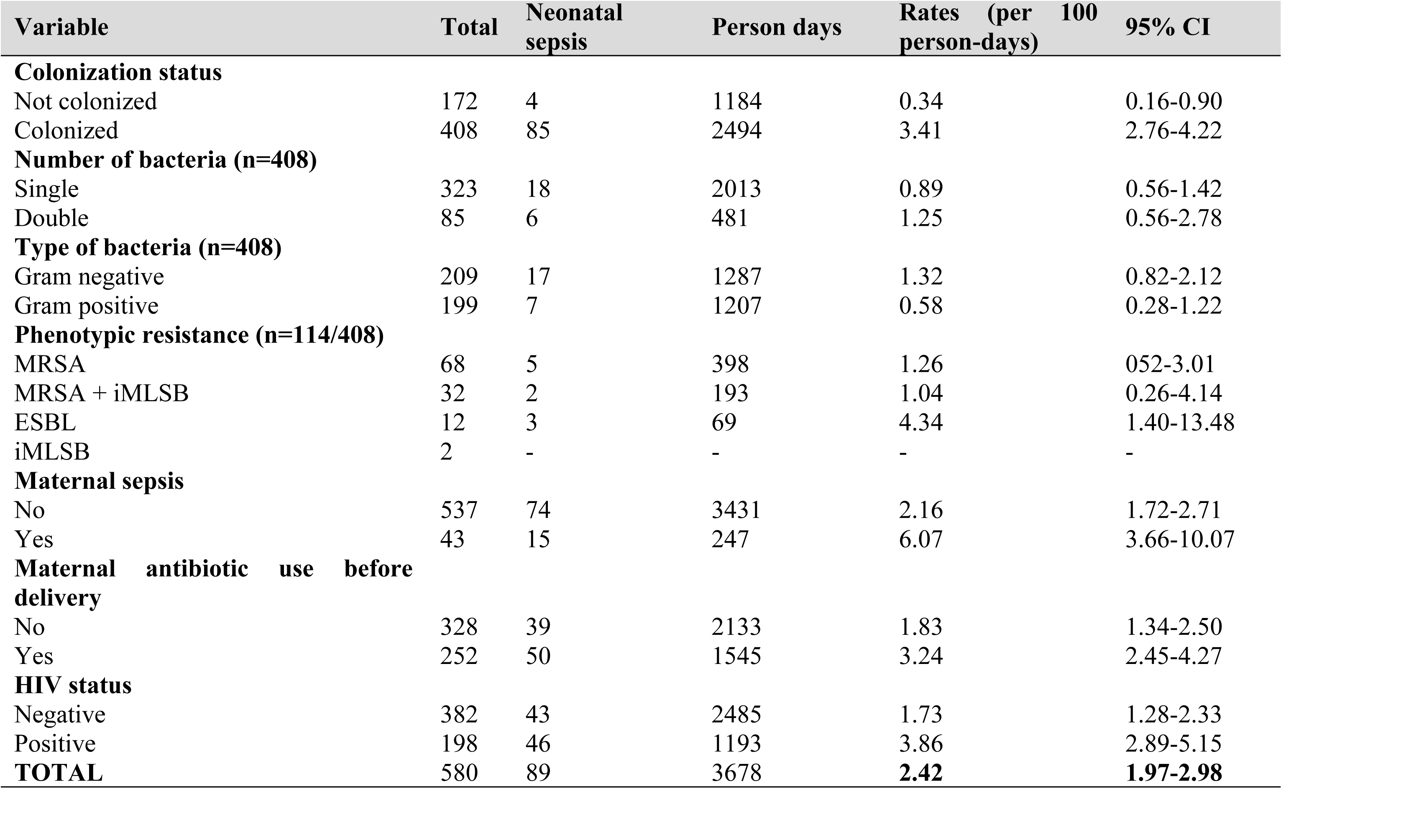
Incidence of Neonatal Sepsis by Clinical and Maternal Characteristics.

Kaplan–Meier survival curves demonstrated differences in neonatal sepsis-free survival according to maternal colonization status, maternal sepsis, and maternal HIV status (Fig 1). Neonates born to colonized mothers had a markedly lower probability of remaining free of neonatal sepsis compared with those born to non-colonized mothers. Similarly, neonates of mothers with maternal sepsis experienced an earlier onset of neonatal sepsis compared with those without maternal sepsis. Neonates born to PWLHIV also had a lower sepsis-free survival probability than those born to HUPW.

**Figure 1.**
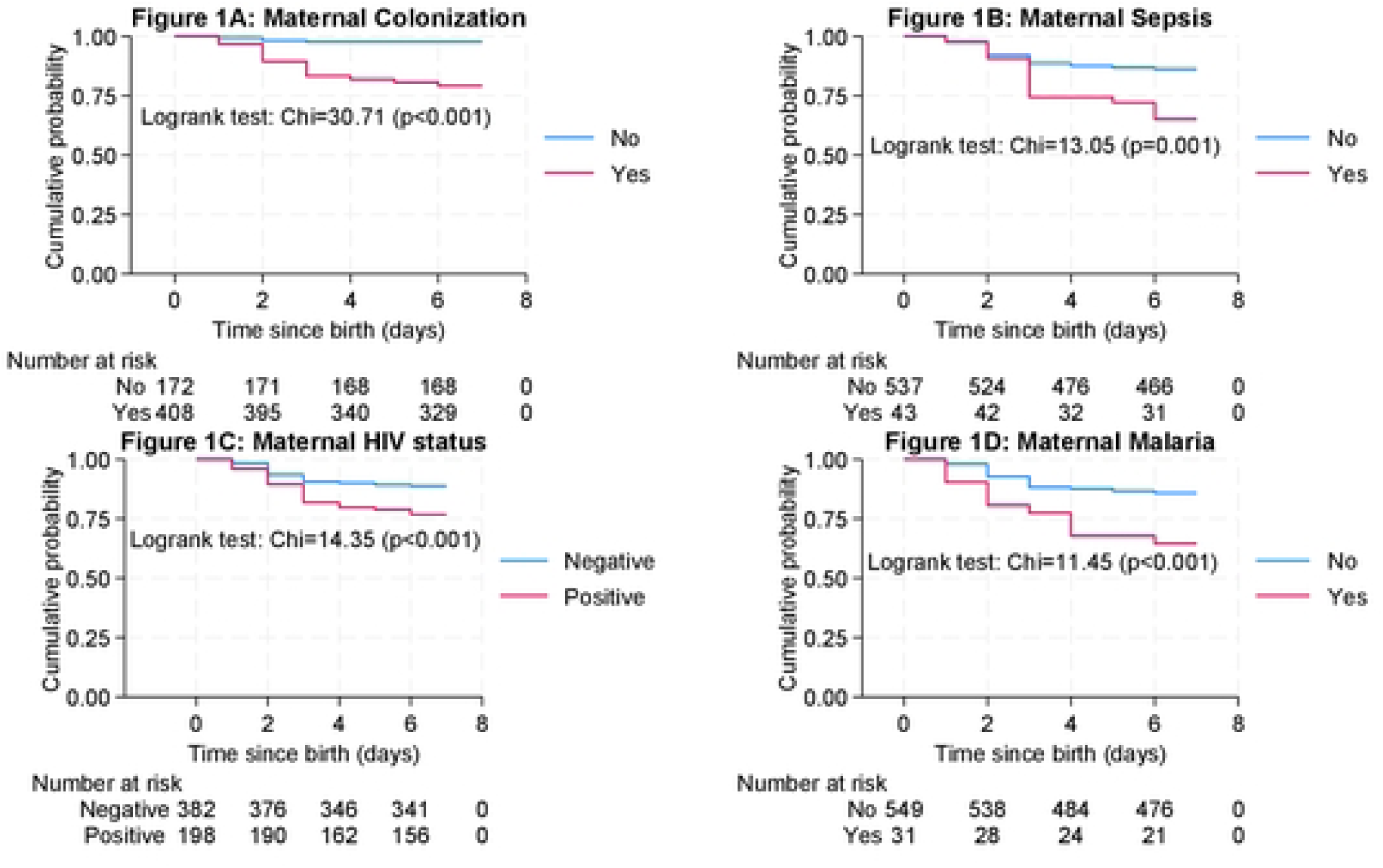
Kaplan—Meier curves of neonatal sepsis-free survival according to maternal colonization, meternal sepsis, and maternal HIV status.

### Proportional Hazard Assumption

In assessing the proportional hazards (PH) assumption, the overall model diagnosis indicated no violation (χ²=9.08, p=0.24), suggesting the PH assumption was generally satisfied (S5 Table). Furthermore, colonization status (p=0.31) and HIV status (p=0.39) both met the assumption. However, maternal sepsis (p=0.04) violated the model assumption (Fig 2).

**Figure 2.**
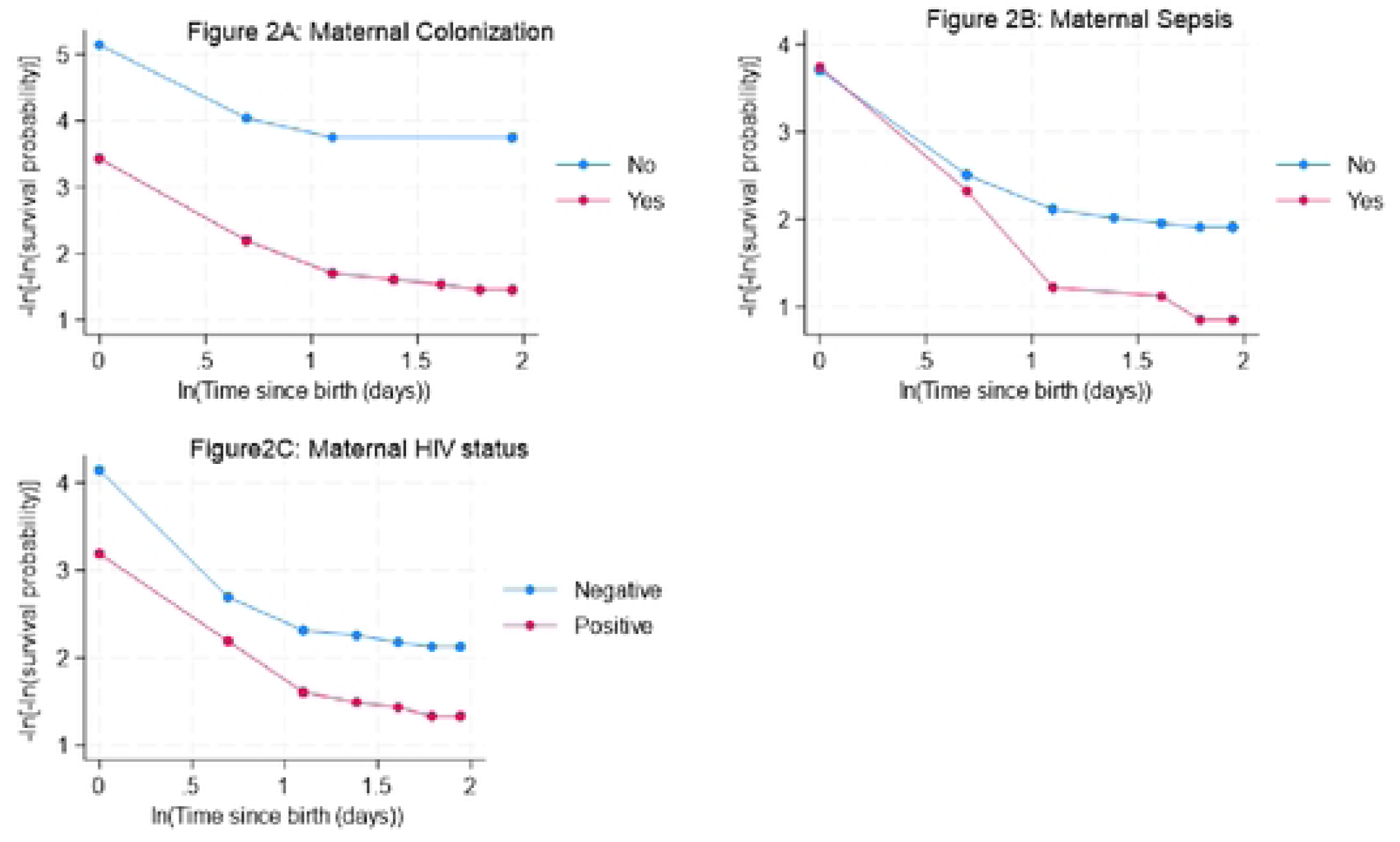
Proportional hazard showing distribution of neonatal sepsis-free by maternal colonization, sepsis, and HIV status.

**Table 5:** Proportional Hazard Assumption.

| Variables | rho | chi | p-value |
| --- | --- | --- | --- |
| Wash underpants with disinfectant | -0.13201 | 1.57 | 0.21 |
| Malaria | 0.01463 | 0.02 | 0.88 |
| Duration of labour | 0.01286 | 0.02 | 0.90 |
| Birth-weight | -0.07025 | 0.45 | 0.50 |
| HIV status | 0.08572 | 0.71 | 0.39 |
| Colonization status | 0.10730 | 1.04 | 0.31 |
| Maternal sepsis | 0.22181 | **4.43** | **0.04** |
| **Overall model** |  | **9.08** | **0.24** |

Abbreviations: *chi*=Chi-square value; *Rho*=residual hazard coefficient; *p-value*=probability value

### Maternal and Neonatal factors associated with neonatal sepsis

In the multivariable Cox proportional hazards model, three variables remained independently associated with neonatal sepsis. Maternal malaria at enrolment increased the hazard of neonatal sepsis nearly threefold (adjusted HR 2.74, 95% CI: 1.43–5.22, p=0.002). Neonates born to PWLHIV had a 76% higher hazard of neonatal sepsis compared with those born to HUPW (adjusted HR 1.76, 95% CI: 1.14–2.73, p=0.011). Maternal colonization was the strongest predictor, with a nearly tenfold increased hazard of neonatal sepsis (adjusted HR 9.52, 95% CI: 3.47–26.09, p=< 0.001) (S6 Table).

**Table 6:** Maternal and Neonatal factors associated with neonatal sepsis.

| Variables | Unadjusted HR |  | Adjusted HR |  |
| --- | --- | --- | --- | --- |
|  | HR, 95% CI | p-value | HR, 95% CI | p-value |
| **Age (in years)** |  |  |  |  |
| 17-24 | 1 |  |  |  |
| 25-35 | 1.11, 0.65-1.92 | 0.69 |  |  |
| 36-44 | 1.21, 0.61-2.39 | 0.59 |  |  |
| **Wash underpants with disinfectant** |  |  |  |  |
| No | 1 |  | 1 |  |
| Yes | 1.69, 0.88-3.27 | 0.12 | 1.57, 0.81-3.06 | 0.18 |
| **PROM** |  |  |  |  |
| No | 1 |  |  |  |
| Yes | 1.49, 0.79-2.81 | 0.21 |  |  |
| **Anaemia** |  |  |  |  |
| No | 1 |  |  |  |
| Yes | 1.30, 0.86-1.99 | 0.22 |  |  |
| **Malaria** |  |  |  |  |
| No | 1 |  | 1 |  |
| Yes | 2.78, 1.48-5.23 | 0.001 | **2.74, 1.43-5.22** | **0.002** |
| **UTIs** |  |  |  |  |
| No | 1 |  |  |  |
| Yes | 1.43, 0.90-2.26 | 0.13 |  |  |
| **Preeclampsia** |  |  |  |  |
| No | 1 |  |  |  |
| Yes | 1.60, 0.87-2.94 | 0.13 |  |  |
| **Duration of labour** | 0.97, 0.92-1.02 | 0.18 | 0.96, 0.92-1.10 | 0.07 |
| **Mode of delivery** |  |  |  |  |
| Spontaneous vaginal delivery | 1 |  |  |  |
| Caesarean section | 1.42, 0.93-2.18 | 0.12 |  |  |
| **Birth-weight** |  |  |  |  |
| Normal birthweight ($\geq 2.5$kg) | 1 | | 1 | |
| Low birthweight ($< 2.5$kg) | 1.93, 1.17-3.21 | 0.001 | 1.54, 0.91-2.62 | 0.11 |
| **Gestation age at birth** |  |  |  |  |
| Post-term (Above 42 weeks) | 0.72, 0.18-2.94 | 0.65 |  |  |
| Term (37-42 weeks) | 1 |  |  |  |
| Premature ($< 37$ weeks) | 1.48, 0.88-2.49 | 0.14 | | |
| **HIV status** |  |  |  |  |
| Negative | 1 |  | 1 |  |
| Positive | 2.15, 1.42-3.27 | $< 0.001$ | **1.76, 1.14-2.73** | **0.01** |
| **Colonization status** |  |  |  |  |
| Not colonized | 1 |  | 1 |  |
| Colonized | 9.63, 3.53-26.25 | $< 0.001$ | **9.52, 3.47-26.09** | **$< 0.001$** |
| **Maternal antibiotic use before delivery** |  |  |  |  |
| No | 1 |  |  |  |
| Yes | 1.73, 1.14-2.63 | 0.010 |  |  |

Among the 24 positive cultures, *E. coli* was the most common isolate (58.3%), followed by *K. pneumoniae* (20.8%) and *S. aureus* (20.7%). Gram-negative organisms showed moderate susceptibility to ceftriaxone and amoxicillin–clavulanic acid, while *S. aureus* isolates were fully susceptible to clindamycin and vancomycin.

There were no major differences in the frequencies or susceptibility patterns of the bacteria, with E. coli, the most common bacterium, being less responsive to treatment in HIV-exposed infants (70-72.7%) while highly susceptible (80-100%) in HIV-unexposed neonates.

## DISCUSSION

In this study, it was found that maternal vaginal bacterial colonization was independently associated with an increased risk of early-onset neonatal sepsis. Notably, maternal colonization was the strongest predictor, conferring a nearly 10-fold increased hazard of neonatal sepsis, and most isolates showed phenotypic concordance consistent with possible maternal–neonatal transmission of pathogenic bacteria during labour and delivery (7). The results differ from a study in Gambia, which found a low prevalence of possible maternal–neonatal transmission (23). These findings offer important insights into how maternal microbiological and clinical factors interact to shape neonatal infection risk. Importantly, this study identified maternal vaginal colonization as a potentially modifiable risk factor. Interventions such as targeted screening, intrapartum antibiotic prophylaxis, or microbiome-based strategies could substantially reduce the burden of neonatal sepsis. However, further investigation is needed to assess the feasibility and cost-effectiveness of such approaches in LMICs. The strong association between maternal vaginal bacterial colonization and neonatal sepsis in this study, together with the high proportion of phenotypically concordant maternal–neonatal isolates, supports the hypothesis that maternal genital tract colonization may contribute to neonatal infection. However, molecular strain typing is needed to establish direct vertical transmission.

The observed incidence of neonatal sepsis in this study is approximately 2.4 cases per 100 person-days. This is higher than the incidence of 0.1-0.5 per 100 neonates in developed countries, but lower than the 4.9–17.0 cases per 100 person-days reported in other studies from LMICs (24). The markedly higher incidence in this study among neonates born to colonized mothers than in those born to non-colonized mothers underscores the critical role of maternal–neonatal transmission in early-onset sepsis pathogenesis and the biological plausibility of maternal–neonatal microbial transmission. These findings are similar to a study conducted in Ethiopia, which showed maternal genital tract colonization as a key reservoir for neonatal pathogens, particularly Gram-negative organisms such as *E. coli* and *K. pneumoniae* (7, 25); but differ from a study in Gambia, which found low prevalence of maternal–neonatal transmission and lack of maternal linkage(23). The large association observed in our study further supports targeted screening and prevention strategies aimed at maternal colonization. Again, neonatal sepsis was numerically more frequent following Caesarean than spontaneous vaginal delivery, although the difference was not statistically significant. This may reflect gut microbiome alterations, as cesarean-born infants miss initial exposure to maternal vaginal and perineal flora. Instead, their early microbial colonization often features environmental hospital bacteria or opportunistic pathogens rather than protective strains (25, 26).

Maternal HIV infection was independently associated with a 76% increased hazard of neonatal sepsis, even after adjusting for potential confounders. The observed association may be consistent with previously proposed mechanisms of increased infectious susceptibility among HIV-exposed infants (27); however, these mechanisms were not directly evaluated in the present study. Importantly, all PWLHIV in this cohort were on ARTs, suggesting that residual risk persists despite treatment and highlighting the need for additional preventive strategies in this population. Potential mechanisms include impaired transplacental transfer of protective antibodies, immune dysregulation, and alterations in maternal and neonatal microbiota (10).

Maternal malaria at enrolment was further a significant predictor of neonatal sepsis, with nearly a threefold increased risk. This association may reflect the broader impact of malaria on maternal immune function, placental integrity, and fetal development, leading to preterm birth, LBW, and small-for-gestational-age babies, which can further predispose to neonatal sepsis (25, 28, 29). The co-existence of malaria in this setting underscores the importance of integrated antenatal care approaches that address multiple overlapping risk factors and management of infectious diseases besides HIV.

Gram-negative organisms predominated among neonatal isolates, with *E. coli* accounting for more than half of the cases. This pattern is consistent with findings from other studies in LMICs and has important implications for empirical antibiotic therapy (7). The observed antimicrobial susceptibility patterns indicate low-to-moderate sensitivity to WHO Access group drugs, with relatively preserved susceptibility to WHO Watch group drugs, such as piperacillin–tazobactam and meropenem. However, resistant phenotypes, including extended-spectrum beta-lactamase (ESBL)-producing organisms and methicillin-resistant *S. aureus* (MRSA), raise concerns about AMR emergence in neonatal pathogens (10, 20, 30). These findings highlight the need for ongoing surveillance and antimicrobial stewardship programs.

Key strengths include a prospective cohort design with longitudinal follow-up, which strengthens the assessment of temporality between maternal exposures and neonatal outcomes. The relatively large sample size also enhances statistical power and generalizability within similar settings. However, key limitations include the inability to measure viral suppression via viral load testing and reliance on clinical criteria to diagnose neonatal sepsis, with limited microbiological confirmation, which may have led to misclassification. Finally, the small number of culture-positive neonates limits the generation of antibiograms to inform specific antimicrobial therapeutic guidelines, underscoring the need to extend future studies in the context of ongoing nationwide AMR surveillance.

## CONCLUSION

Maternal vaginal pathogenic bacterial colonization is independently associated with early-onset neonatal sepsis in this setting, with maternal HIV infection and malaria contributing additional risk. These findings highlight the need for targeted maternal screening and preventive strategies to interrupt vertical transmission pathways and reduce neonatal morbidity and mortality in Mwanza City, Tanzania, and in similar settings elsewhere. Additionally, strengthening microbiological diagnostic capacity and antimicrobial stewardship is essential to address the growing challenge of antimicrobial resistance in neonatal sepsis.

Future research should assess the effectiveness of maternal screening and prophylactic interventions, and explore the underlying immunological and microbiome-related mechanisms linking maternal exposures to neonatal outcomes.

## Data Availability

Data from this study are attached in the supplementary section of this article.

## Acknowledgments

The authors acknowledge support from members of the Obstetrics & Gynecology and Microbiology & Immunology Departments at BMC and CUHAS, as well as all study participants across all study sites.

## ABBREVIATIONS

ANC: Antenatal care; AMR: Antimicrobial Resistance; ART: Antiretroviral therapy; BMC: Bugando Medical Centre; BHC: Buzuruga Health Centre; CLSI: Clinical and Laboratory Standards Institute; CTC: Care & Treatment Centre; CUHAS: Catholic University of Health & Allied Sciences; HUPW: HIV-uninfected Pregnant Women; HIV: Human Immunodeficiency Virus; IHC: Igoma Health Centre; LBW: Low birthweight; L&D: Labour & Delivery; NDH: Nyamagana District Hospital; LMICs: Low-Middle Income Countries; PWLHIV: Pregnant Women Living With HIV; SSA: Sub-Saharan Africa; STRRH: Sekou Toure Regional referral Hospital.

## Availability of data and materials

Data from this study (S2 Appendix) are attached in the supplementary section of this article.

## Author’s contributions

DM, BK, MM, and JS: conceived, designed, and executed the study; DM, GWK, and MJ: participated in data and sample collection; DM, PAS, JD, JS, and BK: were involved in data analysis and interpretation; JD, BK, MM, and JS: supervised the execution of the study; DM: prepared the first manuscript, which was critically reviewed and approved by all authors.

## Supporting information

Fig 1: Kaplan-Meier survival analysis

Fig 2: Proportional hazard

S1 Appendix: Data collection tool

S2 Appendix: VagCol-neonatal Sepsis dataset

